# Clinical performance evaluation of SARS-CoV-2 rapid antigen testing in point of care usage in comparison to RT-qPCR

**DOI:** 10.1101/2021.03.27.21253966

**Authors:** Isabell Wagenhäuser, Kerstin Knies, Vera Rauschenberger, Michael Eisenmann, Miriam McDonogh, Nils Petri, Oliver Andres, Sven Flemming, Micha Gawlik, Michael Papsdorf, Regina Taurines, Hartmut Böhm, Johannes Forster, Dirk Weismann, Benedikt Weißbrich, Lars Dölken, Johannes Liese, Oliver Kurzai, Ulrich Vogel, Manuel Krone

## Abstract

**Background:** Antigen rapid diagnostic tests (RDT) for SARS-CoV-2 are fast, broadly available, and inexpensive. Despite this, reliable clinical performance data is sparse.

**Methods:** In a prospective performance evaluation study, RDT from three manufacturers (NADAL®, Panbio™, MEDsan®) were compared to quantitative reverse transcription polymerase chain reaction (RT-qPCR) in 5 068 oropharyngeal swabs for detection of SARS-CoV-2 in a hospital setting. Viral load was derived from standardized RT-qPCR Cycle threshold (C_t_) values. The data collection period ranged from November 12, 2020 to February 28, 2021.

**Findings:** Overall, sensitivity of RDT compared to RT-qPCR was 42·57% (95% CI 33·38%–52·31%), and specificity 99·68% (95% CI 99·48%–99·80%). Sensitivity declined with decreasing viral load from 100% in samples with a deduced viral load of ≥10^8^ SARS-CoV-2 RNA copies per ml to 8·82% in samples with a viral load lower than 10^4^ SARS-CoV-2 RNA copies per ml. No significant differences in sensitivity or specificity could be observed between the three manufacturers, or between samples with and without spike protein variant B.1.1.7. The NPV in the study cohort was 98·84%; the PPV in persons with typical COVID-19 symptoms was 97·37%, and 28·57% in persons without or with atypical symptoms.

**Interpretation:** RDT are a reliable method to diagnose SARS-CoV-2 infection in persons with high viral load. RDT are a valuable addition to RT-qPCR testing, as they reliably detect infectious persons with high viral loads before RT-qPCR results are available.

**Funding:** German Federal Ministry for Education and Science (BMBF), Free State of Bavaria

**Research in context:** *Evidence before this study:* We searched PubMED an MedRxiv for articles including “COVID-19”, “COVID”, “SARS-CoV-2”, “coronavirus” as well as “antigen detection”, “rapid antigen test”, “Point-of-Care test” in title or abstract, published between January 1, 2020 and February 28, 2021. The more than 150 RDT on the market at the end of February 2021 represent a huge expansion of diagnostic possibilities.^1^ Performance of currently available RDT is evaluated in several international studies, with heterogeneous results. Sensitivity values of RDT range from 0·0%^2^ to 98·3%^3^, specificity from 19·4%^4^ to 100·0%.^2,5–14^. Some of this data differs greatly from manufacturers’ data. However, these previously published performance evaluation studies were conducted under laboratory conditions using frozen swabs, or in small cohorts with middle-aged participants. Comparable RDT performance data from large-scale clinical usage is missing.^5–19^

*Added value of this study:* Based on previous examinations the real life opportunities and limitations of SARS-CoV-2 RDT as an instrument of hospital infection detection and control are still unclear as well as further study results are limited in transferability to general public. Our findings show that RDT performance in daily clinical routine is reliable in persons with high viral for punctual detection and isolation of infectious persons before RT-qPCR become available. In persons with lower viral load, or in case of asymptomatic patients SARS-CoV2 detection by RDT was unsuccessful. The general sensitivity of 42·57% is too low to accept the RDT in clinical use as an alternative to RT-qPCR in diagnosis of COVID-19. Calculated specificity was 99.68%. The results are based on a huge study cohort with more than 5 000 participants including a representative ages structure with pediatric patients up to geriatric individuals, which portrays approximately the demographic structure of the local society.

*Implications of all the available evidence:* Due to the low general sensitivity RDT in clinical use cannot be accepted as an alternative but as an addition to RT-qPCR in SARS-CoV-2 diagnosis. The benefit of early detection of highly infectious persons has to be seen in context of the effort of testing and isolation of false positive tested persons.

## Introduction

For more than a year, the COVID-19 pandemic has been a worldwide public health challenge. As well as contact tracing, contact reduction, quarantine,^20^ and vaccination,^21^ the early testing und detection of infectious persons is key in mitigating the spread of disease.^22^

Due to its high sensitivity and specificity, quantitative reverse transcription polymerase chain reaction (RT-qPCR) has served as the gold standard in diagnosing SARS-CoV-2 since the beginning of the pandemic. However, because these tests require a diagnostic laboratory and more than an hour to complete, they are quite costly, and their availability is limited.^23^

Antigen rapid diagnostic tests (RDT), technically carried out as lateral flow enzyme-linked immunosorbent assays, have become a widely used alternative to RT-qPCR in SARS-CoV-2 diagnostics.^24^ RDT persuade through their point-of-care feasibility, short analysis time, and affordability.^25^

This prospective performance evaluation study compares the accuracy of RDT in comparison to RT-qPCR in daily clinical routine, with a main emphasis on sensitivity in highly infectious individuals and specificity in broad screening use.

## Methods

### Study setting

The study was performed in a 1 438-bed tertiary care hospital in the district of Lower Franconia, Bavaria, Germany. Data collection period ranged from November 12, 2020 to February 28, 2021.

RDT and RT-qPCR SARS-CoV-2 testing was carried out in tandem in key situations to prevent SARS-CoV-2 outbreaks in the hospital. Patients were tested on admission to the medical, pediatric, child, and adolescent psychiatric wards, the surgical emergency department, as well as the delivery room. During the study period, usage of RDT on admission was extended to all other clinical departments of the hospital. Patients and persons accompanying underage patients were tested equally. Employees were tested in case of respiratory symptoms, and after close contacts to SARS-CoV-2 positive persons.

SARS-CoV-2 samples were collected with oropharyngeal swabs for RDT and RT-qPCR by trained medical staff. We did not use nasopharyngeal swabs as they (i) were perceived as being more unpleasant compared to oropharyngeal swabs, (ii) have been associated with serious complications,^26^ and (iii) do not provide advantage with regard to viral load at sampling site.^27^

Data was collected during the second wave of the COVID-19 pandemic in Germany.^28^ In the hospital’s catchment area of Lower Franconia, the average weekly incidence during the study period was 119.21 per 100 000 inhabitants. The maximum of daily new infections was reported on December 23, 2020. Due to a stricter lockdown, case numbers declined in January 2021.^29,30^

### Data collection

RDT, RT-qPCR results, and demographic data were documented in the local hospital information system (HIS) SAP ERP 6.0 (SAP, Walldorf, Germany). Persons were categorized by symptoms into patients with typical COVID-19 symptoms according to comparable COVID-19 case definition of the CDC^31^ and the ECDC^32^ (e.g. fever, dry cough, shortness of breath, new olfactory or taste disorder), and persons without or with atypical symptoms which could be attributed to COVID-19 (e.g. deterioration of general condition, falls, diarrhea). Secondary infections caused by persons tested false negative by RDT were detected using a search of the hospitals’ infection control database.

### Antigen rapid diagnostic tests (RDT)

RDT from three manufacturers were selected by manufacturers’ specifications and availability out of 23 products listed by the German Federal Institute for Drugs and Medical Devices in October 2020:^33^

I. NADAL® COVID-19 Ag Test (Nal von Minden GmbH, Regensburg, Germany)
II. PANBIO™ COVID-19 Ag Rapid Test (Abbott Laboratories, Abbott Park IL, USA)
III. MEDsan® SARS-Cov-2 Antigen Rapid Test (MEDsan GmbH, Hamburg, Germany)

All RDT included in the study target the nucleoprotein antigen of SARS-CoV-2 according to the test manuals. Sensitivities of the RDT are said to range from 92·5% (MEDsan®, no Cycle threshold (C_t_) value specified) over 93·3% (PANBIO™, no C_t_ specified) to 97·6% (NADAL®, C_t_ value 20–30). Specificities are stated as 99·4% (PANBIO™), 99·8% (MEDsan®), or >99·9% (NADAL®).

The PANBIO™ RDT has been evaluated in several studies,^5–16,19^ and reported sensitivity values range from 44·6%^13^ to 91·7%^7^. The specificity was continuously in the range of 98·9%^7^ to 100%.^6,8,9,14^ Three small laboratory or cohort studies are published on NADAL®.^16–18^ Overall sensitivity ranged between 24·3%^17^ and 73·1%^18^, and test specificity estimated at more than 99%.^16–18^ MEDsan® RDT has so far only been assessed in a preprint analysis, with a sensitivity of 45·8%, and a specificity of 97·0%.^13^ This data differs considerably from that provided by the manufacturer.

Two of the three tests (NADAL® and MEDsan®) were approved for use on oropharyngeal swabs. The PANBIO™ RDT is approved for nasopharyngeal swabs only but was used in oropharyngeal swabs in comparison to RT-qPCR for this study. The chosen RDT were distributed to clinical sites depending on availability. All swabs were taken oropharyngeally and processed according to manufacturers’ instructions.

In case of more than one documented RDT per person per day, only the first RDT was included in the study. RDT on test persons with a recent COVID-19 infection and subsequent deisolation were excluded. This category of persons is likely no longer infectious despite persistent RT-qPCR positivity.^34^

### Quantitative reverse transcription polymerase chain reaction (RT-qPCR)

Primary RT-qPCR was carried out in the hospital’s virological diagnostic laboratory using different RT-qPCR methods, performed according to the manufacturers’ instructions. Viral load was determined by retesting all samples to ascertain standardized C_t_ values on MagNA Pure 96 (nucleic acid purification) and the 7500 Real-Time PCR System using FTD SARS-CoV-2 Assay. The following formula was used to calculate viral load of the sample:

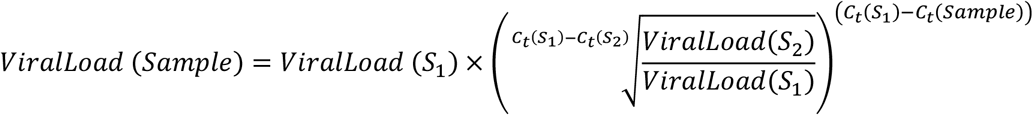

Standards of 10^6^ (S_1_) and 10^7^ (S_2_) SARS-CoV-2 RNA copies per ml were tested three times and resulted in average C_t_ values of 21·3 (S_1_) and 18·2 (S_2_). In two samples with high C_t_ values (34·3 and 37·2) on NeuMoDx™, not enough material was available for retesting, so they were excluded from viral load analysis.

Starting on February 3, 2021, all new RT-qPCR positive samples with sufficient viral load underwent melting curve analysis to detect mutation N501Y, followed by a Δ69-70 deletion PCR to detect variant B.1.1.7. If the mutation N501Y without a Δ69-70 deletion was detected, genome sequencing was performed to detect other variants of concern.

### Ethical approval

The Ethics committee of the University of Wuerzburg waived the need to formally apply for ethical clearance due to the study design (File No. 20210112 01).

### Statistics

Data analysis was performed using Excel® 2019 (Microsoft, Redmond WA, USA), SPSS Statistics 26 (IBM, Armonk NY, USA), and GraphPad Prism (GraphPad Software, San Diego CA, USA).

The Wilson/Brown method was used for confidence interval calculation.^35^ Test performance regarding spike protein variants and symptomatology was compared using Fisher’s exact test. Test performance between manufacturers was compared using Chi-squared test. Viral load between RDT positive and negative persons, as well as between symptomatic, asymptomatic, or atypically symptomatic persons were compared using Mann-Whitney U test. Influence of age on test sensitivity and specificity was analyzed by binary logistic regression. A Pearson correlation coefficient was used to assess the correlation between age and viral load. The two-tailed significance level α was set to 0·05.

### Role of the funding source

This study was initiated by the investigators. The sponsoring institutions had no function in study design, data collection, analysis, and interpretation of data as well as in writing of the manuscript. All authors had unlimited access to all data. The first and the corresponding author had final responsibility for the decision to submit for publication.

## Results

### Test enrollment

Between November 12, 2020 and February 28, 2021, a total of 5 171 parallel RDT and RT-qPCR were carried out. 96 tests were excluded as only the first RDT of each person each day was included. Seven tests were excluded because of persistently positive RT-qPCR results. 5 068 RDT carried out on 4 623 individuals were enrolled and included in the study. NADAL® was used in 810 (15·9%), PANBIO™ in 1 030 (20·36%) and MEDsan® in 3 228 (63·7%) tests (<u>Fig. 1</u>).

**Fig. 1:**
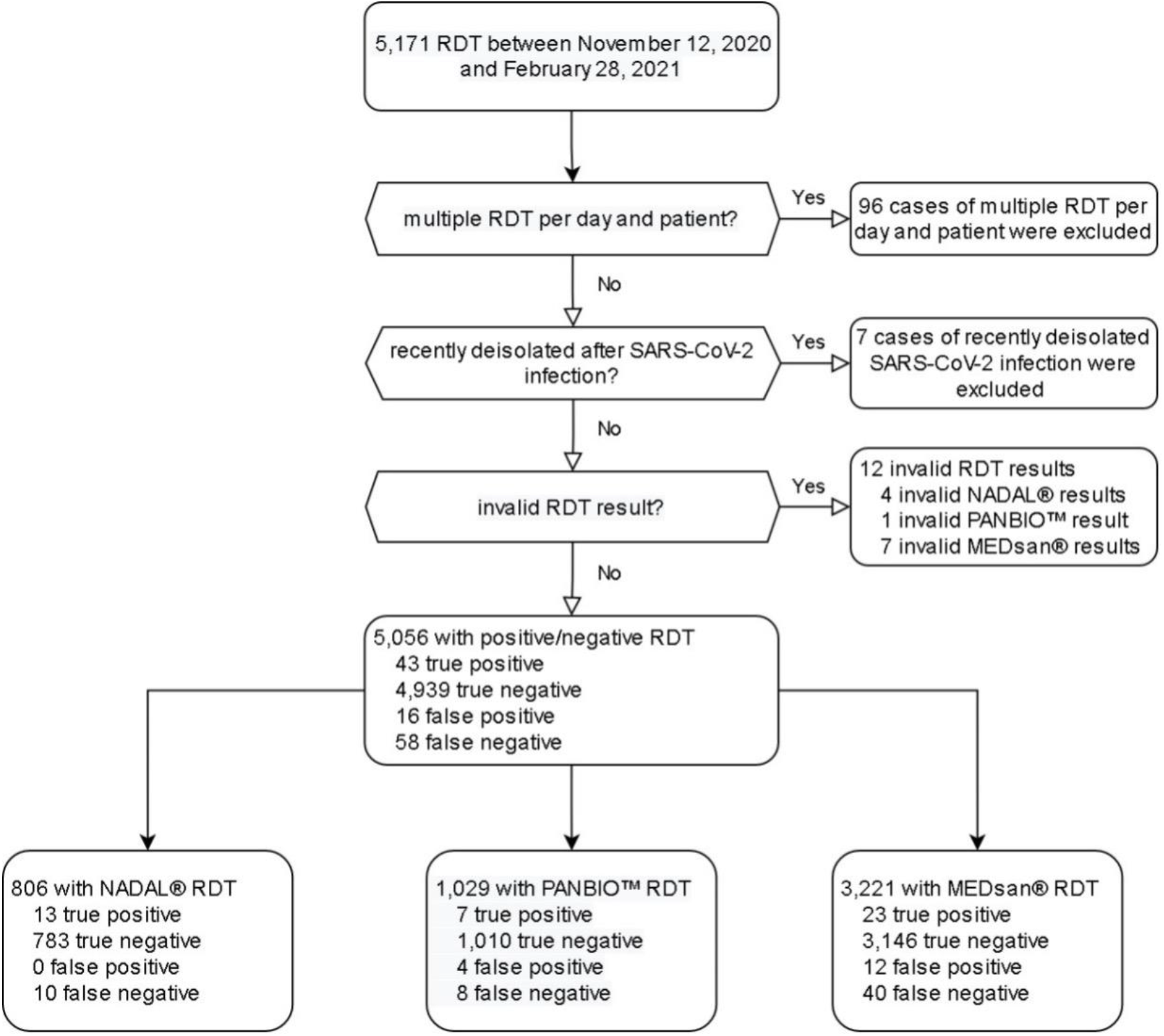
Enrollment of antigen rapid diagnostic test results in the study RDT: Antigen rapid diagnostic test RT-qPCR: Quantitative reverse transcription polymerase chain reaction

### Study population

The tested persons were between 0 and 100 years old (median age: 43 years). 2 677 tests (52·82%) were performed on female, 2 390 (47·16%) on male persons. One test was performed on a person assigned to a diverse gender (0·02%). 4 115 tests were performed on patients (81·20%), 615 on accompanying persons (12·13%), and 338 on staff (6·67%).

Fig. 2 compares the demographics of the study population to the general population. 22·10% of all tested persons were younger than 20 years, 9·41% were 80 years, or older.

**Fig. 2:**
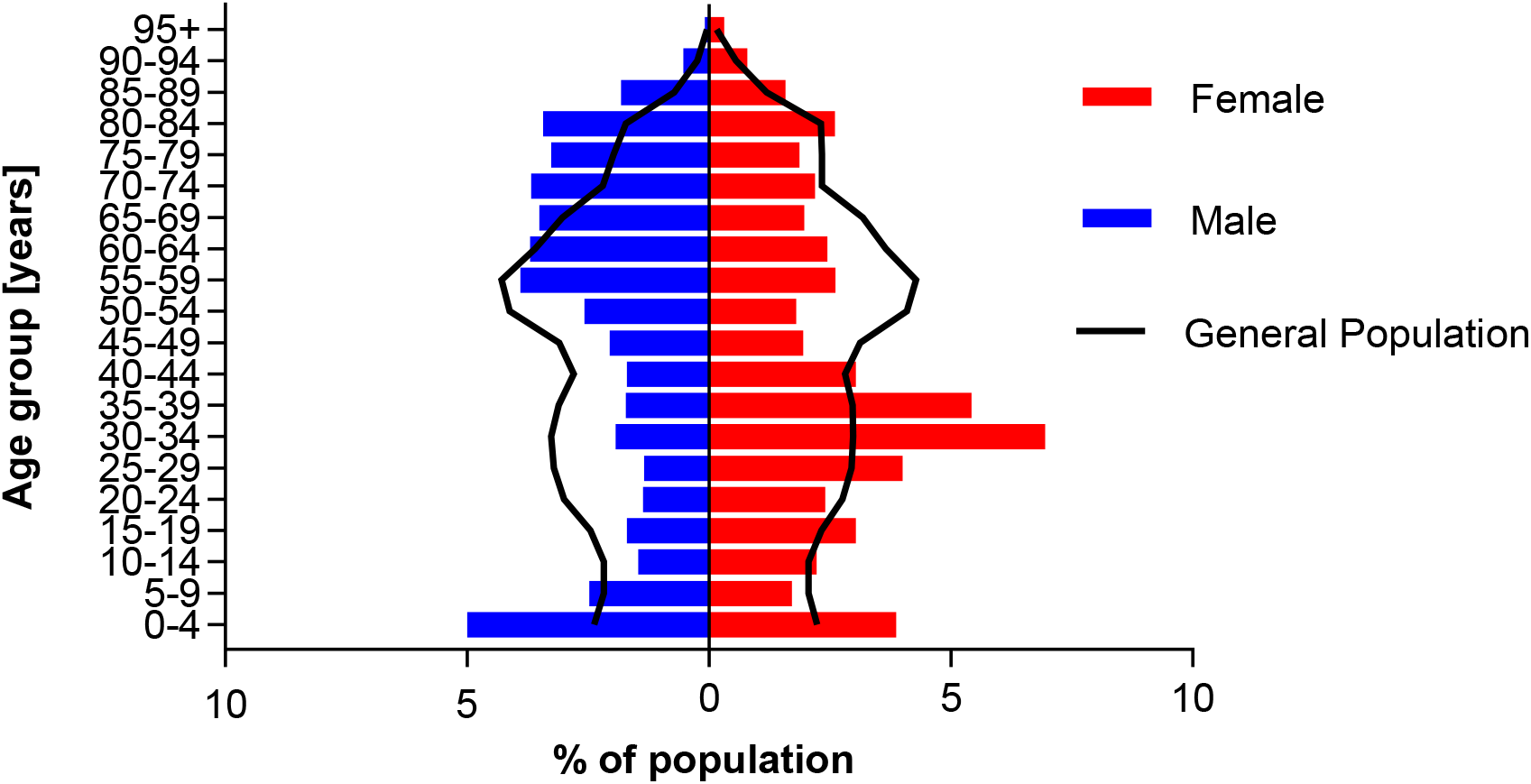
Demographics of the study population compared to the general population of the hospital’s catchment area Study population (red and blue bars, n = 5,067) was compared to the general population of the hospital’s catchment area Lower Franconia (black line, n = 1,317,619) as of December 31, 2019. Due to privacy reasons, one person with diverse gender was excluded from the figure. No data on population with diverse gender was available for Lower Franconia. Population data were obtained from Bavarian federal office for statistics^29^

**Fig. 3:**
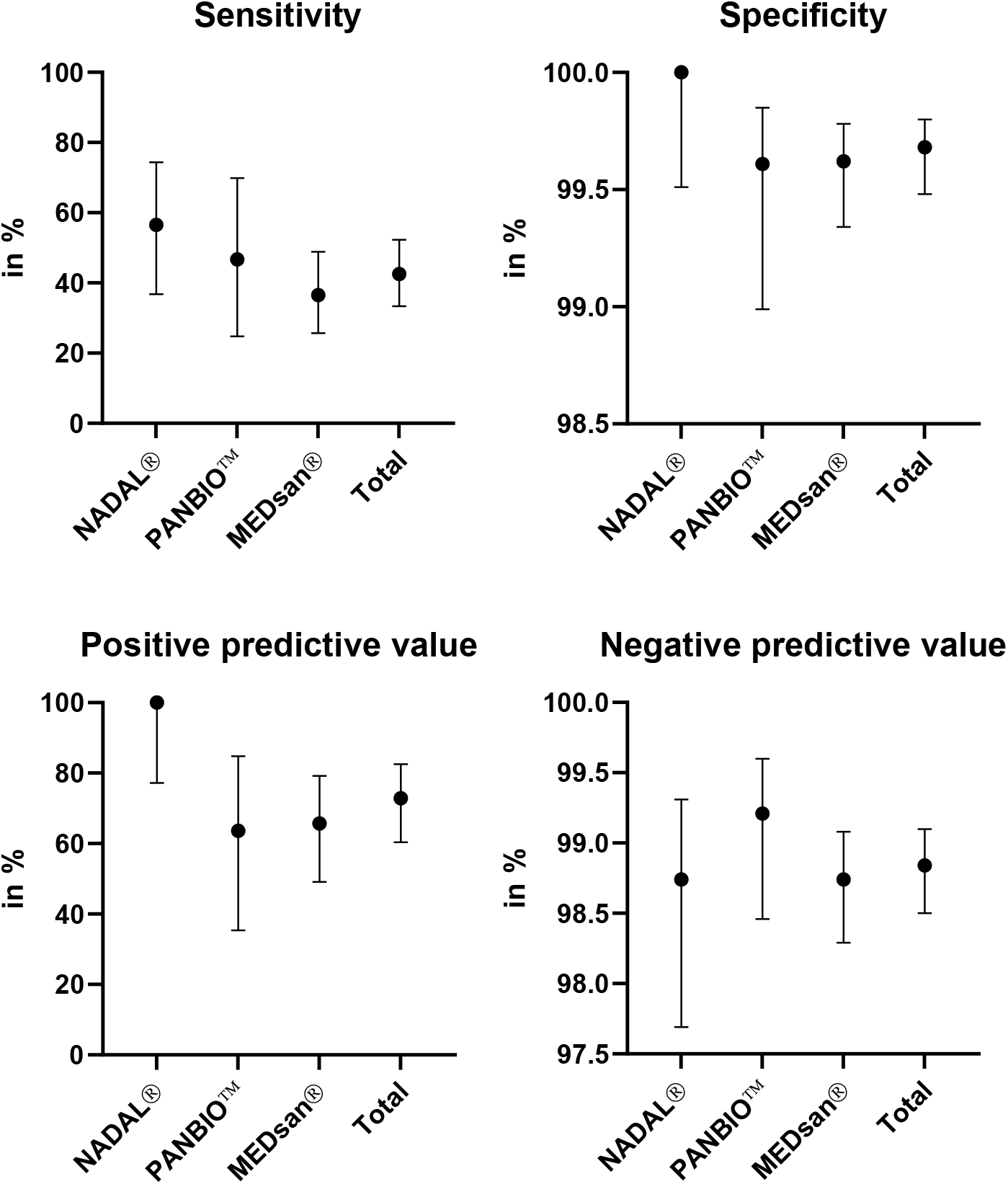
Antigen rapid diagnostic test performance compared to quantitative reverse transcription polymerase chain reaction by manufacturer Sensitivity, specificity, positive predictive value and negative predictive value of antigen rapid diagnostic tests from three manufacturers (nal von minden NADAL®, Abbott PANBIO™, MEDsan®) in comparison to quantitative reverse transcription polymerase chain reaction, n = 5,046

**Fig. 4:**
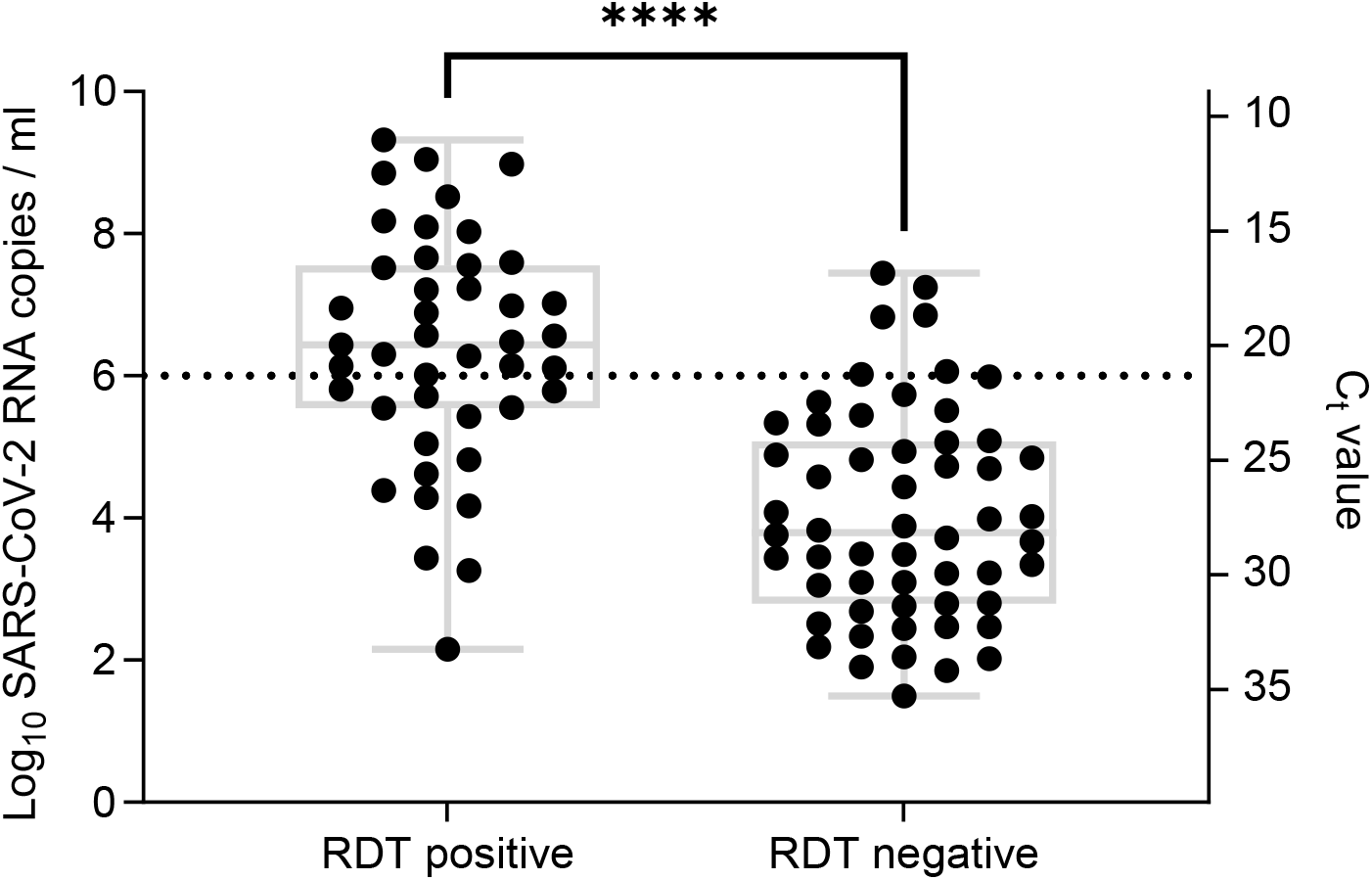
Antigen rapid diagnostic test result in comparison to viral load Viral load was determined by quantitative reverse transcription polymerase chain reaction (RT-qPCR, n=99), dotted line: viral load of 10^6^ SARS-CoV-2 RNA copies per ml. Due to limited sample volume two samples could not be retested using the reference RT-qPCR method. ****: p<0·0001 RDT: Antigen rapid diagnostic test C_t_: Cycle threshold

### Performance of RDT in comparison to RT-qPCR

Out of 5,056 analyzed RDT/RT-qPCR pairs, 101 samples (2·00%) tested positive by RT-qPCR, 59 (1·17%) by RDT. Thus, 43 samples (0·85%) were assessed true positive, 4 939 true negative (97·69%), 16 false positive (0·32%), and 58 false negative (1·15%). Twelve RDT samples were excluded from performance analysis because of their invalid RDT result (negative in the positive control, or interfering lines, 4 NADAL®, 1 PANBIO™, 7 MEDsan®, Fig. 1). Of these, three were RT-qPCR positive.

The overall sensitivity of RDT was 42·57% (95% CI 33·38%–52·31%), the specificity 99·68% (95%CI 99·48%– 99·80%). The positive predictive value (PPV) was 72·88% (95% CI 60·40%–82·56%), and the negative predictive value (NPV) 98·84% (95% CI 98·50%–99·10%).

### Comparison of manufacturers

Sensitivity ranged from 36·51% (23/63, 95% CI, 25·72%–48·18%) for MEDsan® over 46·67% (7/15, 95% CI, 24·81% to 69·88%) for PANBIO™ to 56·52% (13/23, 95% CI 36·81%–74·37%) for NADAL®. Specificity ranged from 99·61% (1 010/1 014, 95% CI 98·99%–99·85%) for PANBIO™ over 99·62% (3 146/3 158, 95% CI 99·34%– 99·78%) for MEDsan® to 100·00% (783/783, 95% CI 99·51%–100·00%) for NADAL®. Differences in sensitivity (p=0·24), and specificity (p=0·22) were not significant (**Fehler! Verweisquelle konnte nicht gefunden werden**.).

### Relation to viral load

C_t_ values in 99 samples tested on the reference system ranged from 11·01 to 35·25 (mean 24·22; SD 5·97), calculated viral loads from 3·16×10^1^ to 2·09×10^9^ SARS-CoV-2 RNA copies per ml. Viral loads in RDT positive persons (median viral load, 2·73 ×10^6^ copies per ml; range, 1·44×10^2^ to 2.09×10^9^) were significantly higher compared to RDT negative persons (median viral load, 6·23×10^3^ copies per ml; range, 3·16×10^1^ to 2·77×10^7^, p<0·0001, **Fehler! Verweisquelle konnte nicht gefunden werden**.).

Sensitivity was 100% in samples with a viral load of ≥10^8^ SARS-CoV-2 RNA copies per ml (8/8, 95% CI 67·56%– 100.00%), 76·92% in samples with a viral load: 10^6^ to 10^8^ copies per ml (20/26, 95% CI 57·95%–88·97%), 38·71% in samples with a viral load of 10^4^ to 10^6^ copies per ml (12/31, 95% CI 23·73%–56·18%), and 8·82% (3/34, 95% CI 3·05%–22·96%) in samples with a viral load <10^4^ copies per ml (Fig. 5).

**Fig. 5:**
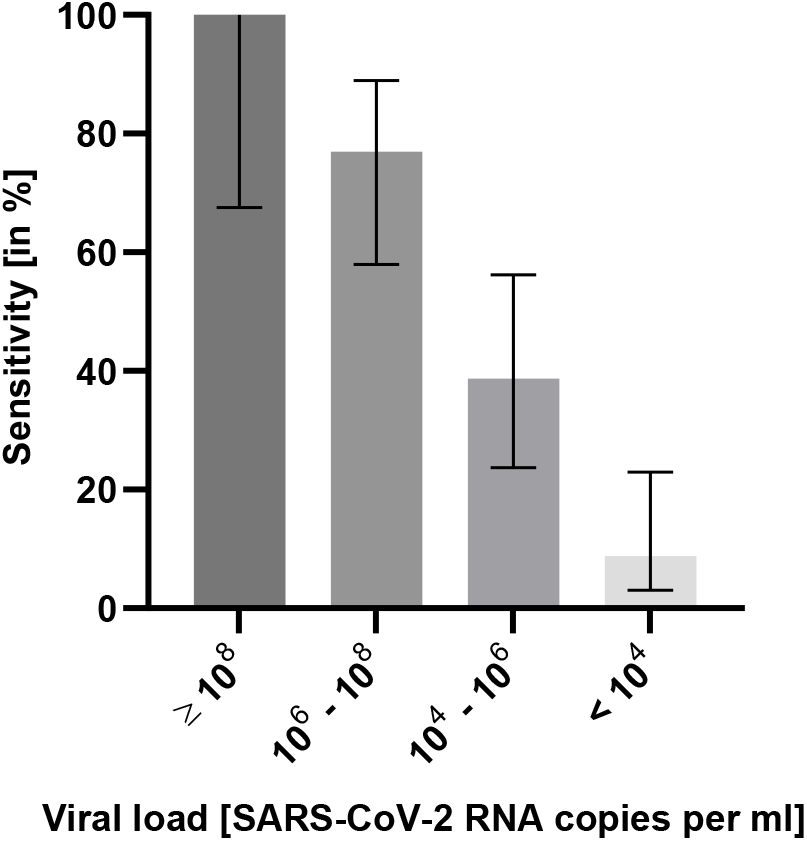
Sensitivity of antigen rapid diagnostic testing in relation to viral load Viral load was determined by quantitative reverse transcription polymerase chain reaction (n=99)

### Relation to spike protein variant

Twenty-three samples were analyzed for a N501Y mutation: ten of these (43·47%) showed a mutation as well as a Δ69-70 deletion compatible with variant B.1.1.7. No other spike protein variants were found. RDT sensitivity (40·00%, 4/10, 95% CI 16·82%–68·73%) did not differ from wild type samples, and samples not analyzed for N501Y mutation (p=1·00).

### Relation to age

RDT sensitivity was lowest in persons <20 years (14·29%, 1/7, 95% CI 0·73%–51·31%), and increased with age to 59·26% (16/27, 95% CI 40·73%–75·49%) in persons ≥80 years. Sensitivity correlated positively with age (p=0·02) as did logarithmical viral load (Correlation coefficient ρ=0·235; p=0·02). There was no significant influence of age on test specificity (ρ=0·010; p=0·03).

### Relation to symptoms

Twenty-five of 101 RT-qPCR positive tests (24·75%) were performed on asymptomatic persons and persons with atypical symptoms which may be attributed to COVID-19, and 76 (75·24%) on persons with typical COVID-19 symptoms. Sensitivity (24·00%, 6/25, 95% CI 11·50%–43·43%), and PPV (28·57%, 6/21, 95% CI 13·81%– 49·96%) were significantly lower in asymptomatic and atypically symptomatic persons compared to persons with typical COVID-19 symptoms. These showed a sensitivity of 48·68% (37/76, 95% CI 37·78%–59·71%, p=0·04), and a PPV of 97·37% (37/38, 95% CI 86·51%–99·87%, p<0·0001). This is in line with higher viral loads in typically symptomatic persons (median: 2·10×10^5^ SARS-CoV-2 RNA copies per ml) compared to asymptomatic or atypically symptomatic persons (median: 9·63×10^3^ copies per ml, p=0·22).

### Secondary infections

One secondary infection was detected a patient who was placed in a two-bed room with an asymptomatic patient after a false negative RDT result (viral load: 6·70×10^6^ SARS-CoV-2 RNA copies per ml).

## Discussion

Our study proves that combining RDT with an RT-qPCR-based test strategy is useful for early detection of persons with high viral load to quickly identify und isolate highly infectious persons before RT-qPCR results are available.

The overall RDT sensitivity of 42·6% differs dramatically from the manufacturers’ information of all three RDT, and is comparable with the results of other publications.^13,14,17,19^ Specificity for all used RDT was above 99·6%, which is comparable to manufacturers’ data as well as performance in other studies.^6–10,13–17,19^ Our data confirms that sensitivity of RDT strongly depends on viral load. Although sensitivity is less than 10% in samples with a low viral load, it reaches 100% with a viral load of more than 10^8^ SARS-CoV-2 RNA copies per ml. As the latter defines potential super-spreaders, it is crucial to identify those individuals as quickly as possible to prevent hospital outbreaks.^36^ The low sensitivity of RDT in persons with low viral loads means these tests must be combined with RT-qPCR. Persons may have a low viral load, and not be infectious, at the end of a previously undiagnosed COVID-19 infection. This load decreases further.^34^ In contrast, viral load at the beginning of a SARS-CoV-2 infection is low, and rapidly increases after the test is performed. Unless these individuals are identified by a parallel RT-qPCR, a false negative RDT may cause and fuel outbreaks.^37^ Additionally, incorrect swabbing may strongly decrease in-vitro viral load in the sample and falsely suggest a lower viral load.^38^ Because they are more susceptible to false negatives with low viral loads, RDT are more prone to sampling problems.

As the PPV is highly dependent on prevalence in the tested population, false positive RDT results do not pose a relevant threat in populations with high prevalence. However, broad use of RDT in asymptomatic individuals in a low prevalence setting may result in a large number of false positive results.

No significant differences in RDT sensitivity or specificity were found between the three products tested. This is especially important because while NADAL® and MEDsan® RDT are approved for both nasopharyngeal and oropharyngeal specimens, PANBIO™ is only approved for nasopharyngeal specimens. Our data indicates that PANBIO™ is comparable the other two RDT in oropharyngeal specimen sampling, which may be better tolerated by patients.

RT-qPCR was positive in three of twelve persons with a documented invalid RDT result. This suggests that persons with atypical lines and thus invalid RDT results should be treated as RDT positive until RT-qPCR results are available.

No differences were found in RDT performance regarding spike protein variant B.1.1.7. This is significant because the proportion of this variant is dramatically increasing worldwide.^39^

Sensitivity was lowest in persons younger than 20 years, as was viral load determined by RT-qPCR. This may represent in-vivo viral load and indicate decreased infectiousness. It may also be explained by the fact that correct sampling procedure in children is more challenging than in adults. This must be considered when RDT is used in children.

Our study has several limitations. For each participant was assessed only by one of the three chosen RDT, and therefore different RDT only compared indirectly. Moreover, the distribution of the three RDT was inhomogeneous throughout the different clinical departments. Each of these also has an individual patient structure. Despite this, our data represents in vivo experience with RDTs in a large cohort. The low incidence of SARS-CoV-2 in our study setting limits the number of RT-qPCR positive persons in the study but reflects a realistic scenario of present and future RDT use. The performance of RDTs in other spike protein variants cannot be assessed as they were not determined in the study population. Given the targets of the assays, however, spike protein mutations are unlikely to affect RDT-detection.

## Conclusion

RDT are a reliable diagnostic tool to quickly detect persons with a high SARS-CoV-2 viral load. Usage of RDT can help to detect and isolate potential super-spreaders before RT-qPCR results are available, especially for persons entering the hospital. RDT can also help to accelerate treatment of critically ill patients by ruling out high infectiousness. However, all used RDT were unsuccessful in detecting persons with lower viral load. This problem may be aggravated by inadequate sampling, and can result in failure to detect patients in an early stage of infection (i.e. with low but rapidly increasing viral loads). Thus, sensitivity of RDT is too low to accept its clinical use as an alternative to RT-qPCR in diagnosing COVID-19 when RT-qPCR is available. In a low incidence scenario, the benefit of detecting highly infectious persons by RDT has to be weighed against the effort of testing and isolating falsely positive tested persons taking into account the SARS-CoV-2 prevalence in the population.

## Data Availability

Individual participant data that underlie the results reported in this article, after de-identification (text, tables, figures, and appendices) as well as the study protocol, statistical analysis plan, and analytic code is made available to researchers who provide a methodologically sound proposal to achieve aims in the approved proposal on request to the corresponding author.

## Author contributions

Ms Wagenhäuser and Dr Krone had full access to all of the data in the study and take responsibility for the integrity of the data and the accuracy of the data analysis.

Concept and design: Andres, Forster, Weismann, Weißbrich, Dölken, Liese, Kurzai, Vogel, Krone. RT-qPCR testing as well as a standardized C_t_ quantification: Knies, Weißbrich.

RDT use and documentation instruction in different departments: Rauschenberger, Eisenmann, Andres, Weismann, Flemming, Gawlik, Papsdorf, Taurines, Böhm, Krone.

User support: Rauschenberger, Eisenmann, Vogel, Krone.

Collection of clinical data from patient’s files: Wagenhäuser, Rauschenberger, Eisenman, McDonogh, Petri, Krone. Statistical analysis: Wagenhäuser, Krone.

Obtained funding: Kurzai, Vogel.

First draft of the manuscript: Wagenhäuser, Krone.

Reviewing and modifying the manuscript and approving its final version: Knies, Rauschenberger, Eisenmann, McDonogh, Petri, Andres, Flemming, Gawlik, Papsdorf, Taurines, Böhm, Forster, Weismann, Weißbrich, Dölken, Liese, Kurzai, Vogel.

## Declaration of competing interest

None of the authors has any conflict of interest.

## Funding

This study was funded by the German Federal Ministry for Education and Science (BMBF) within the program InfectControl (project COVMon, grant-No 03COV26A) and via a grant provided to the University Hospital of Wuerzburg by the Network University Medicine on COVID-19 (B-FAST, grant-No 01KX2021) as well as by the Free State of Bavaria with COVID-research funds provided to the University of Wuerzburg, Germany.

Nils Petri is supported by the German Research Foundation (DFG) funded scholarship UNION CVD.

## Additional contributions

We thank all hospital staff for conducting RDT testing and documenting test results and all laboratory staff in the virological diagnostic laboratory for performing RT-qPCR testing. We thank accounting department from medical controlling for SAP support.

## Notes

### Competing Interest Statement

The authors have declared no competing interest.

## References

1. Foundation of Innovative New Diagnostics: SARS-CoV-2 diagnostic pipeline, https://www.finddx.org/covid%2019/pipeline/?avance=all&type=Rapid+diagnostic+tests&test_target=Antigen&status=all&sect%20ion=show-all&action=default (Accessed 9 March 2021). 2021.

2. Weitzel T, Legarraga P, Iruretagoyena M, et al. Comparative evaluation of four rapid SARS-CoV-2 antigen detection tests using universal transport medium. Travel Med Infect Dis 2020; 39: 101942.

3. Chaimayo C, Kaewnaphan B, Tanlieng N, et al. Rapid SARS-CoV-2 antigen detection assay in comparison with real-time RT-PCR assay for laboratory diagnosis of COVID-19 in Thailand. Virol J 2020; 17(): 177.

4. Schildgen V, Demuth S, Lüsebrink J, Schildgen O. Limits and opportunities of SARS-CoV-2 antigen rapid tests – an experience based perspective. medRxiv 2020: 2020.09.22.20199372.

5. Agullo V, Fernandez-Gonzalez M, Ortiz de la Tabla V, et al. Evaluation of the rapid antigen test Panbio COVID-19 in saliva and nasal swabs in a population-based point-of-care study. J Infect 2020.

6. Albert E, Torres I, Bueno F, et al. Field evaluation of a rapid antigen test (Panbio COVID-19 Ag Rapid Test Device) for COVID-19 diagnosis in primary healthcare centres. Clin Microbiol Infect 2020.

7. Alemany A, Baro B, Ouchi D, et al. Analytical and clinical performance of the panbio COVID-19 antigen-detecting rapid diagnostic test. J Infect 2021.

8. Berger A, Ngo Nsoga MT, Perez-Rodriguez FJ, et al. Diagnostic accuracy of two commercial SARS-CoV-2 Antigen-detecting rapid tests at the point of care in community-based testing centers. medRxiv 2020: 2020.11.20.20235341.

9. Gremmels H, Winkel BMF, Schuurman R, et al. Real-life validation of the Panbio COVID-19 antigen rapid test (Abbott) in community-dwelling subjects with symptoms of potential SARS-CoV-2 infection. EClinicalMedicine 2021; 31: 100677.

10. Krüger LJ, Gaeddert M, Tobian F, et al. Evaluation of the accuracy and ease-of-use of Abbott PanBio - A WHO emergency use listed, rapid, antigen-detecting point-of-care diagnostic test for SARS-CoV-2. medRxiv 2020: 2020.11.27.20239699.

11. Lanser L, Bellmann-Weiler R, Öttl KW, et al. Evaluating the clinical utility and sensitivity of SARS-CoV-2 antigen testing in relation to RT-PCR Ct values. Infection 2020: 1–3.

12. Linares M, Perez-Tanoira R, Carrero A, et al. Panbio antigen rapid test is reliable to diagnose SARS-CoV-2 infection in the first 7 days after the onset of symptoms. J Clin Virol 2020; 133: 104659.

13. Olearo F, Nörz D, Heinrich F, et al. Handling and accuracy of four rapid antigen tests for the diagnosis of SARS-CoV-2 compared to RT-qPCR. medRxiv 2020: 2020.12.05.20244673.

14. Torres I, Poujois S, Albert E, Colomina J, Navarro D. Evaluation of a rapid antigen test (Panbio COVID-19 Ag rapid test device) for SARS-CoV-2 detection in asymptomatic close contacts of COVID-19 patients. Clin Microbiol Infect 2021.

15. Abdulrahman A, Mustafa F, AlAwadhi AI, Alansari Q, AlAlawi B, AlQahtani M. Comparison of SARS-COV-2 nasal antigen test to nasopharyngeal RT-PCR in mildly symptomatic patients. medRxiv 2020: 2020.11.10.20228973.

16. Corman Vmh, V. C.; Bleicker, T.; Schmidt, M. L.; Mühlemann, B.; Zuchowski, M.; Jó Lei, W. K.; Tscheak, P.; Möncke-Buchner, E.; Müller, M. A.; Krumbholz, A.; Drexler, J. F.; Drosten, C. Comparison of seven commercial SARS-CoV-2 rapid Point-of-Care Antigen tests. 2020.

17. Kohmer N, Toptan T, Pallas C, et al. The Comparative Clinical Performance of Four SARS-CoV-2 Rapid Antigen Tests and Their Correlation to Infectivity In Vitro. J Clin Med 2021; 10(2).

18. Stromer A, Rose R, Schafer M, et al. Performance of a Point-of-Care Test for the Rapid Detection of SARS-CoV-2 Antigen. Microorganisms 2020; 9(1).

19. Villaverde S, Dominguez-Rodriguez S, Sabrido G, et al. Diagnostic Accuracy of the Panbio SARS-CoV-2 Antigen Rapid Test Compared with Rt-Pcr Testing of Nasopharyngeal Samples in the Pediatric Population. J Pediatr 2021.

20. Nussbaumer-Streit B, Mayr V, Dobrescu AI, et al. Quarantine alone or in combination with other public health measures to control COVID-19: a rapid review. Cochrane Database Syst Rev 2020; 4(4): Cd013574.

21. Mello MM, Silverman RD, Omer SB. Ensuring Uptake of Vaccines against SARS-CoV-2. New England Journal of Medicine 2020; 383(14): 1296–9.

22. Krüger S, Leskien M, Schuller P, et al. Performance and feasibility of universal PCR admission screening for SARS-CoV-2 in a German tertiary care hospital. J Med Virol 2021.

23. Corman VM, Landt O, Kaiser M, et al. Detection of 2019 novel coronavirus (2019-nCoV) by real-time RT-PCR. Euro Surveill 2020; 25(3).

24. Loeffelholz MJ, Tang YW. Laboratory diagnosis of emerging human coronavirus infections - the state of the art. Emerg Microbes Infect 2020; 9(1): 747–56.

25. Young S, Taylor SN, Cammarata CL, et al. Clinical Evaluation of BD Veritor SARS-CoV-2 Point-of-Care Test Performance Compared to PCR-Based Testing and versus the Sofia 2 SARS Antigen Point-of-Care Test. J Clin Microbiol 2020; 59(1).

26. Sullivan CB, Schwalje AT, Jensen M, et al. Cerebrospinal Fluid Leak After Nasal Swab Testing for Coronavirus Disease 2019. JAMA Otolaryngology–Head & Neck Surgery 2020; 146(12): 1179–81.

27. Wölfel R, Corman VM, Guggemos W, et al. Virological assessment of hospitalized patients with COVID-2019. Nature 2020; 581(7809): 465–9.

28. Robert Koch-Institut: COVID-19-Dashboard, https://experience.arcgis.com/experience/478220a4c454480e823b17327b2bf1d4 (Accessed 4 March 2021).

29. Bayerisches Landesamt für Statistik: Die Gliederung der Bevölkerung nach Kreisen sowie nach Alter, Geschlecht und nach Deutschen und Ausländern, https://www.statistik.bayern.de/mam/produkte/veroffentlichungen/statistische_berichte/a1300c_201800.xla (Accessed 17 February 2021).

30. Bayerisches Landesamt für Gesundheit und Lebensmittelsicherheit: Übersicht der Fallzahlen von Coronavirusinfektionen in Bayern, https://www.lgl.bayern.de/gesundheit/infektionsschutz/infektionskrankheiten_a_z/coronavirus/karte_cor onavirus/ (Accessed 17 February 2021).

31. Centers for Disease Control and Prevention (CDC): Interim Guidelines for Collecting and Handling of Clinical Specimens for COVID-19 Testing, https://www.cdc.gov/coronavirus/2019-ncov/lab/guidelines-clinical-specimens.html (Accessed 3 March 2021).

32. European Centre for Disease Prevention and Control (ECDC): Diagnostic testing and screening for SARS-CoV-2, www.ecdc.europa.eu/en/covid-19/latest-evidence/diagnostic-testing (Accessed 3 March 2021). 2020.

33. Bundesinstitut für Arzneimittel und Medizinprodukte: Antigen-Tests zum direkten Erregernachweis des Coronavirus SARS-CoV-2, https://antigentest.bfarm.de/ords/f?p=101:100:13556100388097:::::&tz=1:00 (Accessed on 23 October 2020).

34. Vibholm LK, Nielsen SSF, Pahus MH, et al. SARS-CoV-2 persistence is associated with antigen-specific CD8 T-cell responses. EBioMedicine 2021; 64.

35. Brown L, Cai, T., & DasGupta, A.. Interval Estimation for a Binomial Proportion, Statist. Sci, 16(2), 101–133; 2001.

36. Avadhanula V, Nicholson EG, Ferlic-Stark L, et al. Viral load of SARS-CoV-2 in adults during the first and second wave of COVID-19 pandemic in Houston, TX: the potential of the super-spreader. J Infect Dis 2021.

37. Krone M, Noffz A, Richter E, Vogel U, Schwab M. Control of a COVID-19 outbreak in a nursing home by general screening and cohort isolation in Germany, March to May 2020. Eurosurveillance 2021; 26(1): 2001365.

38. Richard-Greenblatt M, Ziegler MJ, Bromberg V, et al. Impact of Nasopharyngeal Specimen Quality on SARS-CoV-2 Test Sensitivity. medRxiv 2020.

39. Galloway SE, Paul P, MacCannell DR, et al. Emergence of SARS-CoV-2 B.1.1.7 Lineage - United States, December 29, 2020-January 12, 2021. MMWR Morb Mortal Wkly Rep 2021; 70(3): 95–9.

